# Incorporating county-level NO_2_ data into a metapopulation model: a case study of the first COVID-19 wave in Oklahoma

**DOI:** 10.64898/2026.09.26.26364084

**Authors:** Haridas K. Das, Patrick Stephens, Lucas M. Stolerman

## Abstract

Nitrogen dioxide (NO_2_) is a widespread air pollutant associated with the risk and prevalence of various respiratory infectious diseases. Epidemiological studies have reported associations between NO_2_ exposure and COVID-19 incidence, and biological evidence indicates that NO_2_ exposure increases vulnerability to infection. Motivated by these findings and by the scarcity of mechanistic epidemic models that incorporate air pollutant data, here we introduce an NO_2^-^_ and mobility-informed metapopulation model and study the first COVID-19 wave in Oklahoma. County-level NO_2_ concentrations enter the infection rates through a first-order parameterization, alongside high-resolution mobility patterns capturing heterogeneous inter-county connectivity. Using numerical simulations, we demonstrate that higher environmental sensitivity produces earlier and more synchronized outbreaks across counties, while lower sensitivity yields delayed, asynchronous trajectories. When fitted to county-level COVID-19 incidence data, our model outperformed the classic SIR model (fitted independently in each county) in over 75% of counties in Oklahoma across all standard error metrics. The improvements were even more pronounced in forecasting tasks across epidemic stages, with county-level win rates against the SIR model of 59.74% (surge), 87.01% (peak), 80.52% (post-peak), and 88.31% (decline) by nRMSE. To isolate the contribution of NO_2_, we compared our model against a mobility-only model with NO_2_ removed and homogeneous infection rates across counties. The NO_2_ term improved county-level forecasts during the surge (94.81% of counties by nRMSE), post-peak (88.31%), and peak (62.34%) stages, though not during the early and decline stages, suggesting that NO_2_ information can generate better forecasts than mobility alone during periods of high incidence. Our results demonstrate that environmental exposure can enter the transmission structure of a mechanistic epidemic model directly and that doing so yields measurable forecasting gains.

## 1. Introduction

The COVID-19 pandemic showed the limits of our preparedness to anticipate and respond to emerging infectious diseases (*1, 2*). The challenge of characterizing outbreak dynamics and supporting public health decision-making has propelled the development of novel mechanistic epidemic models to deliver transparent insights, test rigorous policy scenarios, and generate reliable forecasts (*3*). Considerable work has extended classical modeling frameworks to incorporate climate variables (*4, 5*), human mobility (*6, 7*), behavioral adaptation (*8*), and vaccination uptake (*9*). In contrast, chemical exposure drivers such as air pollution have received markedly less attention in mechanistic modeling, despite a substantial body of biological and epidemiological evidence linking ambient pollutants to respiratory disease risk and outcomes (*10–12*). This imbalance leaves a meaningful gap in our ability to capture the environmental dimension of early epidemic dynamics, particularly in regions where pollution exposure varies substantially across short spatial scales.

Among indicators of urban air pollution, nitrogen dioxide (NO_2_) has emerged as particularly relevant for infectious disease transmission. Epidemiological studies have reported associations between NO_2_ exposure and increased COVID-19 incidence, severity, and mortality across multiple countries, including the United States (*13–18*). NO_2_ is emitted primarily from vehicular traffic and industrial combustion, producing pronounced spatial heterogeneity across urban and rural environments. Oklahoma is of particular interest from this perspective because it lies almost entirely within the “lung cancer belt” of the US (*19, 20*) and also shows heavy economic dependence on oil and gas production (*21*). The latter industries are major emission sources for NO_2_ and other gaseous pollutants (*22*). A recent study tested for the influence of four gaseous pollutants, NO_2_, CO, SO_2_, and O_3_, on county-level COVID-19 incidence during the first three years of the pandemic (*23*). The study found that after accounting for the influence of confounding factors such as population density, county area, and the percentage of elderly individuals, NO_2_ showed a positive and near-linear relationship with incidence. The effect was strongest at concentrations above 74 *µ*mol*/m*^2^. Relationships observed for the other three pollutants were weaker and more variable, in some cases no longer showing significant correlations in some models that included all four variables. In contrast, the association between incidence and NO_2_ was extremely robust, persisting in all models regardless of whether other gaseous pollutants were included or excluded.

Biological evidence indicates plausible mechanisms linking exposure to NO_2_ and other air pollutants to COVID-19 transmission. Clear relationships have already been established between NO_2_ exposure, airway inflammation, and vulnerability to respiratory infections (*24–26*). Early in the pandemic, Wang et al. raised the possibility that pollution-driven impairment of human immune responses could raise transmission and thereby aid the spread of SARS-CoV-2 (*27*). Concentrating on NO_2_, O_3_, and particulate matter (PM), Woodby et al. suggested that the oxidative stress these pollutants generate in the lungs could modulate the host proteases that SARS-CoV-2 depends on to enter cells (*28*). More recently, Di Ciaula et al. proposed a more direct link: among 147 patients, higher NO_2_ exposure was accompanied by altered lymphocyte populations and signs of immune dysfunction, leading the authors to argue that such environmentally driven changes may arise before infection rather than as a consequence of it, leaving exposed individuals more vulnerable to COVID-19 (*18*). Such increased vulnerability can be understood as a greater likelihood of acquiring SARS-CoV-2 infection upon exposure or, in modeling terms, as an enhanced per-contact transmission probability. This correspondence opens a clear path for integrating environmental exposure directly as a component of the transmission process. While statistical studies have explored associations between NO_2_ and reproduction numbers (*29, 30*), to our knowledge, no prior study has explicitly incorporated NO_2_ into the transmission rate of a mechanistic epidemic model and assessed its ability to characterize and forecast disease dynamics.

Here we address this gap by incorporating county-level NO_2_ data into a metapopulation SIR-network model (*31, 32*) to describe the first COVID-19 wave in Oklahoma. Leveraging the spatial granularity of satellite-derived NO_2_ measurements, which previous studies have shown are a reliable proxy for ground-level measurements (*33–36*), we couple county-specific infection rates that depend on pre-pandemic NO_2_ levels to a network in which counties are connected through the daily movement of their residents, characterized by mobility data from GPS-enabled devices and summarized as pre- and post-lockdown flux matrices. We evaluate the model against a classic SIR model (*37*) in each county by fitting epidemiological data and forecasting incidence across epidemic stages, with uncertainty quantified via parametric bootstrapping (*38*). Our NO_2^-^_ and mobility-informed model substantially improves the representation of county-level transmission dynamics and yields more accurate forecasts than the SIR model across most counties and epidemic phases. To determine whether these gains reflect the NO_2_ term specifically rather than the network structure alone, we further compare our model against a reduced version in which the NO_2_ contribution is removed. This comparison shows that NO_2_ improves county-level forecasts during periods of high incidence (surge, peak, and post-peak stages). Our work thus demonstrates that air pollution information can improve epidemic forecasting.

## 2. Data

We provide details on county-level reported COVID-19 cases, nitrogen dioxide (NO_2_) concentrations, and mobility data in the state of Oklahoma.

### 2.1 COVID-19 data sources and preprocessing

County-level COVID-19 case data for the United States were obtained from the Center for Systems Science and Engineering (CSSE) at Johns Hopkins University (*39*). For each Oklahoma county, we aggregated reported COVID-19 case counts by epidemiological week from March 2020 to March 2023. For this study, we restricted the analysis to the period from March 2020 to July 2021.

### 2.2 Mobility data

We utilized aggregated SafeGraph data (2020) (*40, 41*), covering roughly 10% of GPS-enabled devices and validated against Google mobility reports (*42*), which have been widely applied in epidemiological studies of COVID-19 transmission (*43–46*). Our study focused on the state of Oklahoma, comprising 77 counties, where county-level mobility networks capture local population flows as a dynamic network: counties serve as nodes, and directed edges represent daily population movements between counties.

### 2.3 Nitrogen dioxide (NO_2_) air pollution data

We incorporated high-resolution satellite-derived NO_2_ concentrations, specifically leveraging observations from the Ozone Monitoring Instrument (OMI) and the Sentinel-5P TROPOMI sensor (*47, 48*). To generate this covariate, we aggregated county-level NO_2_ measurements (mg/m^3^) from the Sentinel-5P TROPOMI instrument. Since biological evidence links vulnerability to respiratory infection to sustained exposure, acting through airway injury and altered immune function, we averaged pre-pandemic levels over the period between January 1 and March 1, 2020, before the first COVID-19 case in Oklahoma. We denote this average as ⟨NO_2_⟩_*j*_, representing the characteristic exposure of county *j* to NO_2_, fixed throughout the study period.

## 3. Methods

In this section, we describe the modeling framework and analytical procedures used in this study. We begin with a brief discussion of the SIR model as a baseline, followed by the construction of the mobility flux matrix and the formulation of environmentally modulated infection rates. Next, we describe the NO_2^-^_ and mobility-informed model, along with our forecasting window partitioning approach, parameter estimation procedures, and the uncertainty quantification framework.

### 3.1 SIR Model

The Susceptible-Infectious-Recovered (SIR) compartmental model is one of the most traditional and widely used frameworks to study the dynamics of infectious diseases and inform public health interventions at the population level (*37*). This SIR model assumes a well-mixed population in which all individuals interact uniformly and is described by the following system of differential equations:

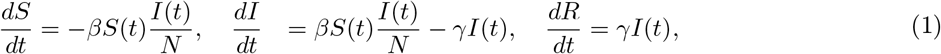

where *S*(*t*), *I*(*t*), *R*(*t*) denote susceptible, infectious, and recovered populations, respectively. Here, *N* = *S*+*I* +*R* is the total population, *β* is the infection rate, and *γ* is the recovery rate.

### 3.2 Construction of origin-destination mobility flux matrices

We constructed a county-level mobility flux matrix to capture heterogeneous mobility patterns. Following our previous work (*46*), we define

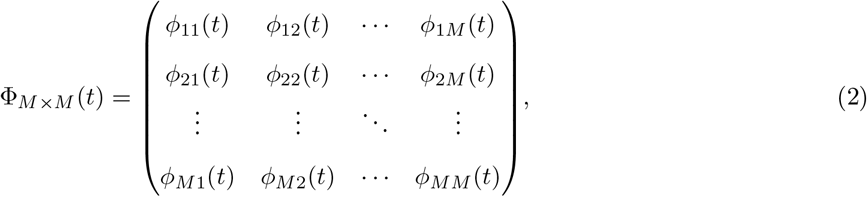

where each element *ϕ*_*ij*_(*t*) represents the normalized fraction of movement from county *i* to county *j*, and *M* = 77 denotes the total number of counties in the state of Oklahoma. Each row of Φ_*M×M*_ (*t*) sums to one; that is, ∑_*j*_ *ϕ*_*ij*_(*t*) = 1.

To incorporate lockdown effects while maintaining computational efficiency, we derived two static mobility flux matrices, Φ^pre^ ∈ R^*M ×M*^ and Φ^post^ ∈ R^*M ×M*^, by averaging the time-dependent mobility fluxes over the pre-lockdown period (January 1, 2020–March 28, 2020) and the post-lockdown period (March 29, 2020–December 31, 2020), respectively (*46*). The mobility matrix used in the model is therefore defined as

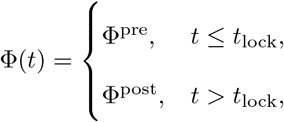

where *t*_lock_ = March 28, 2020 denotes the mobility-transition date associated with statewide COVID-19 restrictions. These matrices capture mean county-to-county mobility rates for each period, and the model switches between them based on the current time step.

### 3.3 NO_2^-^_dependent infection rates

To build NO_2_-dependent infection rates *β*_*j*_, we follow the terminology adopted by Weiss (*49*) and define *κ* as the number of contacts an infected individual has per unit of time. The fraction *τ*_*j*_ of adequate contacts in county *j* is assumed to increase with higher local NO_2_ levels, reflecting the hypothesis that gaseous pollutants influence vulnerability to COVID-19 infections (*18, 27, 28*). We assume the first-order approximation *τ*_*j*_ = *τ*_0_ + *χ*⟨NO_2_⟩_*j*_ where *τ*_0_ is a baseline value and *χ* ≥ 0 is a sensitivity factor small enough that *τ*_*j*_ ≤ 1. Here, ⟨NO_2_⟩_*j*_ represents the average NO_2_ concentration in county *j* between January 1, 2020, and March 1, 2020, a period before the first COVID-19 case in Oklahoma (*50*). The expression for the infection rate is then given by *β*_*j*_ = *κτ*_*j*_, or equivalently,

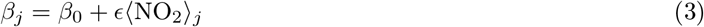

where *β*_0_ = *κτ*_0_ and *ϵ* = *κχ*. Here *β*_0_ is a baseline infection rate and *ϵ* has units of (time^*−*1^ concentration^*−*1^).

### 3.4 NO_2^-^_ and mobility-informed metapopulation model

We employed a metapopulation SIR-network model (*31*) to describe the spread of infectious diseases across interconnected regions. The state of Oklahoma is represented as a network of *M* = 77 counties, where each county corresponds to a node. Here *V* = {1, 2, …, *M* } denotes the set of counties in the network, and the weighted edges represent the fraction *ϕ*_*ij*_(*t*) ∈ [0, 1] of residents moving daily from county *i* to county *j*. The connectivity structure of the network is encoded in the temporal mobility flux matrix Φ_*M×M*_ (*t*) (Eq. 2).

In this framework, the population from each county *i* ∈ *V* is divided into susceptible (*S*_*i*_(*t*)), infectious (*I*_*i*_(*t*), including quarantined or hospitalized individuals) and recovered (*R*_*i*_(*t*)) individuals. The disease dynamics are governed by the following system of differential equations:

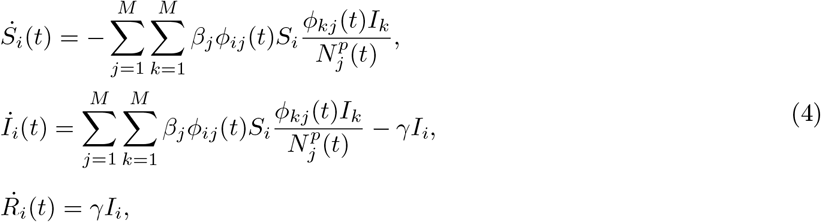

where *N*_*i*_ is the population size and *β*_*i*_ is the infection rate for county *i* given by Eq. 3. Here *γ* is the recovery rate, and the present population at node *i* is given by 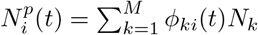.

COVID-19 transmission is well known to be affected by asymptomatic cases, quarantine and isolation measures, exposure periods, loss of immunity, and vaccination coverage (*51–54*). In this work, we adopted the simplified SIR framework to isolate and quantify the combined effects of mobility and an NO_2_-dependent infection rate on county-level disease dynamics. Moreover, since we focus on the first COVID-19 wave in Oklahoma— characterized by a surge in cases in early 2020, epidemic peaks over the winter, and a decline in cases in early spring 2021—the SIR structure provides an adequate characterization for this particular window. A modeling study of COVID-19 transmission spanning wider time windows, and hence periods of new variants such as Delta and Omicron, would require an extended model incorporating loss of immunity terms.

The basic reproduction number R_0_ can be computed as the spectral radius (largest eigenvalue) of the next-generation matrix (NGM) (*55, 56*). Within our framework, this quantity has been previously studied for various network structures (*32*). For a flux matrix with entries *ϕ*_*ij*_, the NGM is given by

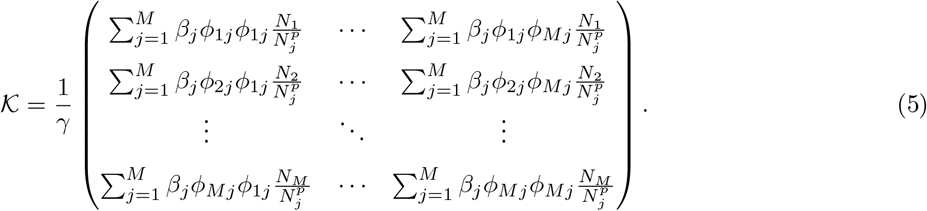

In subsection 4.2, we numerically compute R_0_ using the flux matrix Φ^pre^ and determine the region where R_0_ = 1 by varying the infection rate parameters *β*_0_ and *ϵ* (Eq. 3).

### 3.5 Characterizing epidemic stages of infectious disease dynamics

To account for the different periods in the first COVID-19 wave in Oklahoma, we introduced five epidemic stages (*57–59*): Early, Surge, Peak, Post-peak, and Decline, and examined how the NO_2^-^_ and mobility-informed model captured the real disease dynamics in comparison to the SIR model. Table 1 summarizes the epidemic windows, associated stages, and corresponding fitting and forecast periods used in this study.

**Table 1:** Epidemic stages from March 07, 2020, to July 03, 2021 (the full study period). Each window uses a progressively longer training period, starting with the first observation and ending at the indicated end date.

| Window Name | Start Date | End Date | Forecast Start | Forecast End |
| --- | --- | --- | --- | --- |
| Early | March 07, 2020 | July 18, 2020 | July 25, 2020 | August 22, 2020 |
| Surge | March 07, 2020 | September 26, 2020 | October 03, 2020 | October 31, 2020 |
| Peak | March 07, 2020 | December 04, 2020 | December 11, 2020 | January 08, 2021 |
| Post-peak | March 07, 2020 | February 13, 2021 | February 20, 2021 | March 20, 2021 |
| Decline | March 07, 2020 | April 24, 2021 | May 01, 2021 | May 29, 2021 |

### 3.6 Model parameters and initial conditions

Tables 2 and 3 summarize the model parameters, initial conditions, and whether parameters were fixed or estimated through model fitting in this study. For the standard SIR model (Eq. 1), we estimated the infection rate *β* and the recovery rate *γ*. In contrast, the NO_2^-^_ and mobility-informed model incorporates inter-county mobility data, and the parameters estimated from model fitting were the baseline infection rate *β*_0_, the recovery rate *γ*, and the NO_2_-related sensitivity parameter *ϵ*, which modulates the county-specific infection rates *β*_*j*_ (Eq. 3). For both models, we additionally estimated the reporting rate *ρ* (see parameter estimation details below). Here, we performed all numerical simulations in MATLAB R2024b using the ode45 solver, a fourth/fifth-order Runge–Kutta–Fehlberg method. Additionally, the numerical solutions were computed on a uniform time grid with spacing Δ*t* = 0.1 over a simulation horizon of *T* days (e.g., *T* = 88), i.e., *t* ∈ {0, Δ*t*, …, *T* }.

**Table 2:**
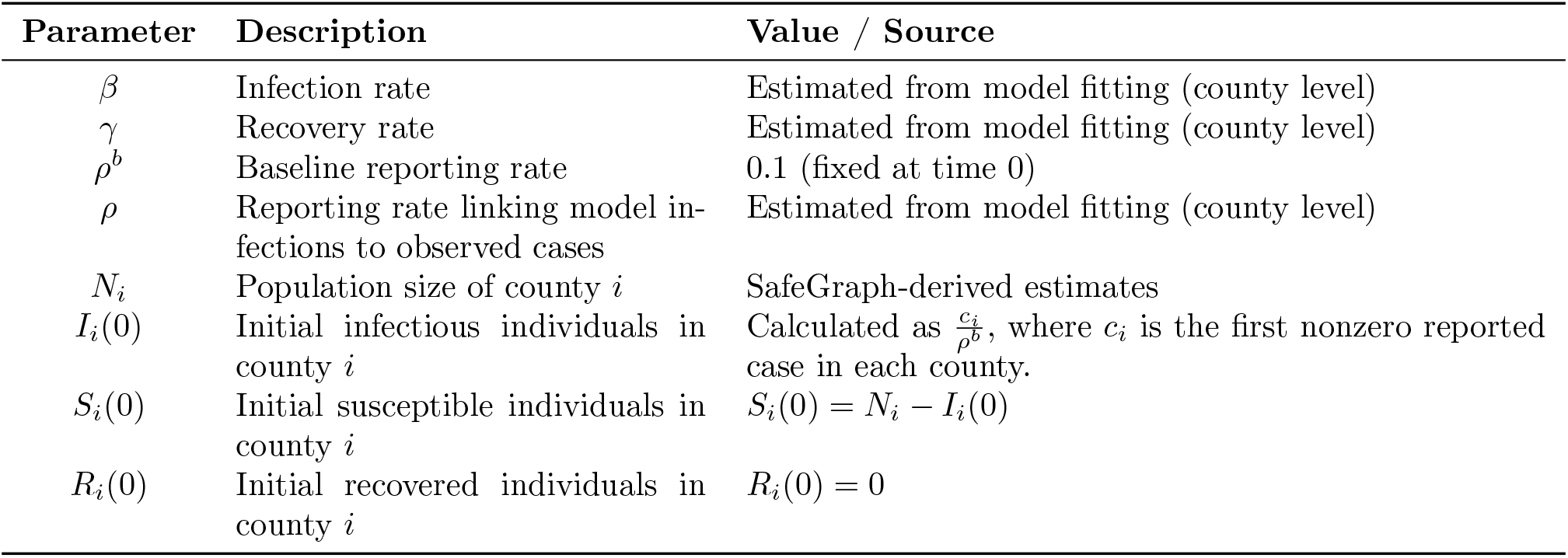
Model parameters and initial conditions for the SIR model.

| Parameter | Description | Value / Source |
| --- | --- | --- |
| $\beta$ | Infection rate | Estimated from model fitting (county level) |
| $\gamma$ | Recovery rate | Estimated from model fitting (county level) |
| $\rho^b$ | Baseline reporting rate | 0.1 (fixed at time 0) |
| $\rho$ | Reporting rate linking model infections to observed cases | Estimated from model fitting (county level) |
| $N_i$ | Population size of county $i$ | SafeGraph-derived estimates |
| $I_i(0)$ | Initial infectious individuals in county $i$ | Calculated as $\frac{c_i}{\rho^b}$, where $c_i$ is the first nonzero reported case in each county. |
| $S_i(0)$ | Initial susceptible individuals in county $i$ | $S_i(0) = N_i - I_i(0)$ |
| $R_i(0)$ | Initial recovered individuals in county $i$ | $R_i(0) = 0$ |

**Table 3:** Model parameters and initial conditions for the NO_2^-^_ and mobility-informed metapopulation model.

| Parameter | Description | Value / Source |
| --- | --- | --- |
| $M$ | Number of counties in the state | $M = 77$ (Oklahoma metapopulation) |
| $\langle \text{NO}_2 \rangle_j$ | Average nitrogen dioxide level in county $j$ | County-level $\text{NO}_2$ averages (satellite-derived) |
| $\beta_0$ | Baseline infection rate | Estimated from model fitting (global parameter) |
| $\epsilon$ | $\text{NO}_2$ sensitivity parameter | Estimated from model fitting (global parameter) |
| $\rho^b$ | Baseline reporting rate | 0.1 (fixed at time 0) |
| $\rho$ | Reporting rate linking model infections to observed cases | Estimated from model fitting (global parameter) |
| $\beta_j$ | Infection rate in county $j$ | $\beta_j = \beta_0 + \epsilon \langle \text{NO}_2 \rangle_j$ |
| $\phi_{ij}(t)$ | Fraction of population traveling from county $i$ to $j$ at time $t$ | Derived from SafeGraph mobility data |
| $N_i$ | Population size of county $i$ | SafeGraph-derived estimates |
| $\gamma$ | Recovery rate | Estimated from model fitting (global parameter) |
| $I_i(0)$ | Initial infectious individuals in county $i$ | $I_{\text{Tulsa}}(0) = 1/\rho^b$; $I_i(0) = 0$ otherwise |
| $S_i(0)$ | Initial susceptible individuals in county $i$ | $S_i(0) = N_i - I_i(0)$ |
| $R_i(0)$ | Initial recovered individuals in county $i$ | $R_i(0) = 0$ |

### 3.7 Optimization procedure for model fitting and forecasting

We estimated the model parameters (see Tables 2 and 3) by minimizing the error between the observed weekly reported cases of COVID-19 and the predicted infections generated by the SIR model (Eq. 1) and the NO_2^-^_ and mobility-informed model (Eq. 4). More specifically, we estimated *β, γ*, and *ρ* for the SIR model, whereas for the NO_2^-^_ and mobility-informed model, we estimated *β*_0_ (Eq. 3), *γ, ϵ*, and *ρ*.

For the optimization procedure, we denote *y*_*t*_ as the observed number of cases in week *t*, and *ŷ*_*t*_(*θ*) the corresponding model-predicted value. Here, *θ* represents the model’s parameter vectors: *θ* = (*β, ρ, γ*) for the SIR model and *θ* = (*β*_0_, *ϵ, γ, ρ*) for the NO_2^-^_ and mobility-informed model. Under the assumption that reported cases represent a fraction of the true infectious population, the model output is given by *ŷ*_*t*_(*θ*) = *ρI*_*t*_(*θ*), where *I*_*t*_(*θ*) is the prevalence. Then, the parameter estimation was performed by minimizing the sum of squared residuals (SSR),

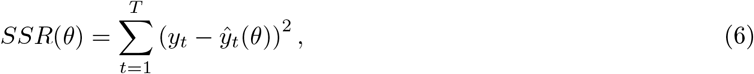

where *T* denotes the total number of weekly observations used for model fitting. The corresponding optimization procedures for the SIR model and NO_2^-^_ and mobility-informed model are described in the supplementary material sections A.2 and A.3, respectively.

To evaluate model performance across different epidemic stages, we implemented an expanding-window fitting strategy coupled with fixed-horizon forecasting. Starting from the first reported case on 07 March 2020, model parameters were sequentially estimated using the progressively longer training windows from Table 1. For each window, five-week-ahead forecasts were generated starting one week after the end of the training period. Our approach provides a basis for evaluating model performance across early, surge, peak, post-peak, and decline phases.

### 3.8 Uncertainty quantification via parametric bootstrapping

To assess the uncertainty in model parameters and resulting epidemic predictions, we applied a parametric bootstrap approach. Observed epidemic incidence data often exhibit overdispersion, where the variance exceeds that expected under a simple Poisson process with mean equal to the variance (*38*). Ignoring overdispersion can lead to overly narrow confidence intervals and an underestimation of uncertainty. To account for this additional variability, we simulated stochastic variability using a negative binomial distribution, which allows extra variance beyond the mean. For week *t*, the simulated incidence was drawn as

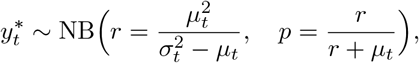

where *µ*_*t*_ and 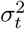 are the mean and variance of the predicted counts, and 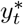 is the simulated observation. Each synthetic dataset was refitted using the original best-fit parameters as initial values, producing an ensemble of parameter estimates and incidence trajectories. Pointwise 95% confidence intervals for predictions were then computed as 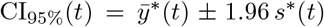, where 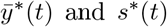 denote the mean and standard deviation across all bootstrapped predictions at time *t*. This approach captures both parameter uncertainty and the additional stochastic variability observed in real-world epidemic data, providing confidence bounds for county-level forecasts across different epidemic stages. The detailed algorithmic steps for parametric bootstrapping in the SIR and NO_2^-^_ and mobility-informed models are provided in the Supplementary Materials (Algorithm S1 and Algorithm S2, respectively).

## 4. Results

Starting from the SIR-network model (*31*), we first developed a framework to incorporate mobility and environmental heterogeneity within the metapopulation, named the NO_2^-^_ and mobility-informed model (see Methods), to account for county-level heterogeneity. We emphasize that the NO_2^-^_ and mobility-informed model introduces variability in disease dynamics through simulation and demonstrates its usefulness for COVID-19 forecasting at different epidemic stages, compared to the classic SIR model (*37*).

Epidemics were initiated by introducing infectious individuals in Tulsa County, obtained from the earliest reported COVID-19 case data (*39*), while all other counties were initially susceptible. Additionally, we used the county population sizes directly from the demographic data (*40, 41*). For each parameter combination (*β*_0_, *ϵ*), the model was simulated forward in time to compute epidemic outcomes, including total epidemic size, peak fraction infected, and outbreak timing.

### 4.1 The first COVID-19 wave in Oklahoma and our modeling approach

We considered our NO_2^-^_ and mobility-informed model as a candidate description of the first COVID-19 wave in Oklahoma (March 2020–July 2021). Data on confirmed COVID-19 cases exhibited substantial spatial and temporal heterogeneity across counties (Fig. 1A). Counties such as Oklahoma and Tulsa experienced rapid early growth, while others, including Cimarron and Latimer, had slower, delayed outbreaks. These differences likely reflect variations in county-level mobility, urbanization, and local environmental factors.

**Figure 1:**
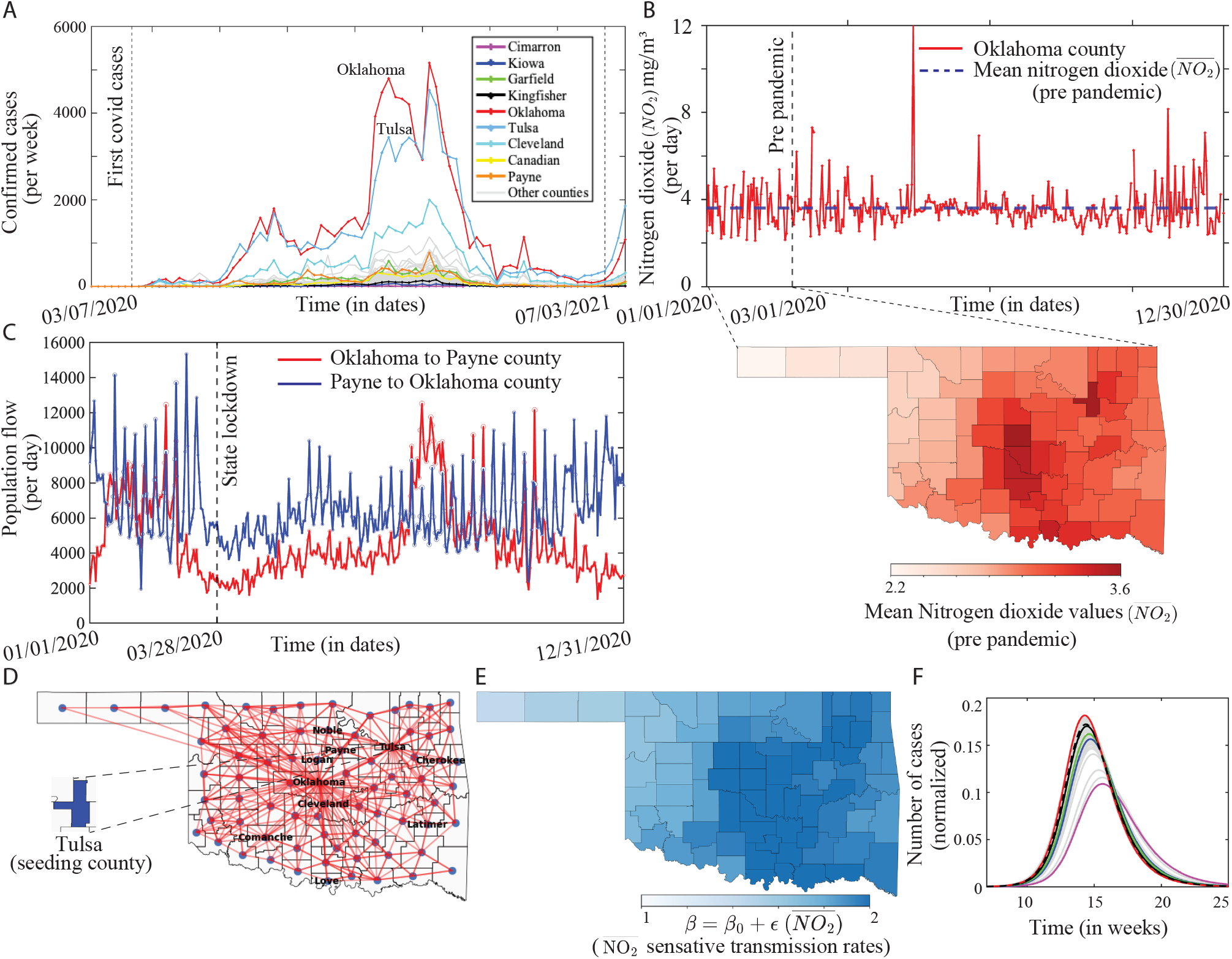
Integrating NO_2_ and mobility data into a metapopulation model: county-level inputs and synthetic simulations for Oklahoma. **A**, Weekly confirmed COVID-19 cases across counties from March 07, 2020 to July 03, 2021; vertical dashed lines indicate the study period of primary interest. **B**, Daily NO_2_ concentrations (mg/m^3^) in Oklahoma County (red line) compared with the pre-pandemic mean 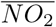 for that county (blue dashed line). The vertical dashed line marks March 1, 2020. Here, the map shows the distribution of pre-pandemic mean 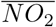 across all counties. **C**, Daily population flow between Oklahoma and Payne counties from January to December 2020; the vertical dashed line indicates the timing of the statewide lockdown. **D**, Network map of inter-county population flows representing the top 10% of population flows (percentile-based threshold), highlighting dominant mobility pathways relevant for disease spread. **E**, Choropleth map of county-level infection rates (*β*) reflecting sensitivity to NO_2_ along the baseline infection rate *β*_0_. **F**, County-level synthetic simulations of the NO_2^-^_ and mobility-informed metapopulation model over time (weeks) from the “seeding county” (Tulsa), showing heterogeneous prevalence patterns across counties.

Motivated by recent studies suggesting that NO_2_ levels often influence variation in realized COVID-19 incidence (*14–18, 60*), and do so within the state of Oklahoma in particular (*23*), we integrated county-specific NO_2_ concentrations at pre-pandemic levels and county-level mobility data to inform our model (see methods). Fig. 1B illustrates variations in NO_2_ concentrations over 2020. The inset shows a map with mean NO_2_ emissions over a pre-pandemic period (Jan 1 – March 1, 2020) for each county in Oklahoma. Fig. 1C exhibits two time series of population flows derived from GPS-enabled mobile devices obtained from SafeGraph (*40*). Fig. 1D depicts the state of Oklahoma as a network, where every county is connected through population flows. Our model integrates mobility and NO_2_-dependent infection rates *β* (Fig. 1E, see methods). Synthetic simulations from the NO_2^-^_ and mobility-informed model can thus produce heterogeneous prevalence patterns across counties (Fig. 1F), consistent with the variability observed in the empirical outbreak data.

### 4.2 Emergent variability in disease trajectories

Variability in disease trajectories can arise from mobility across counties and spatial heterogeneity in transmission. To assess the contribution of NO_2_-dependent infection rates, we examined epidemic outcomes under varying levels of nitrogen dioxide (NO_2_) sensitivity *ϵ* and baseline infection rate *β*_0_ (Eq. 3). Fig. 2 shows four possible trajectories that yield different epidemic sizes and/or state-level peaks, as indicated by the central colormaps. The white dashed line in both colormaps represents the epidemic threshold region where R_0_ = 1. High *ϵ* values lead to early and largely synchronized outbreaks across counties (panels A and B in Fig. 2), whereas lower *ϵ* results in delayed and asynchronous epidemics (panels C and D in Fig. 2). Counties with low baseline NO_2_ levels, such as Cimarron, peak later than counties with high NO_2_ concentrations, such as Oklahoma.

**Figure 2:**
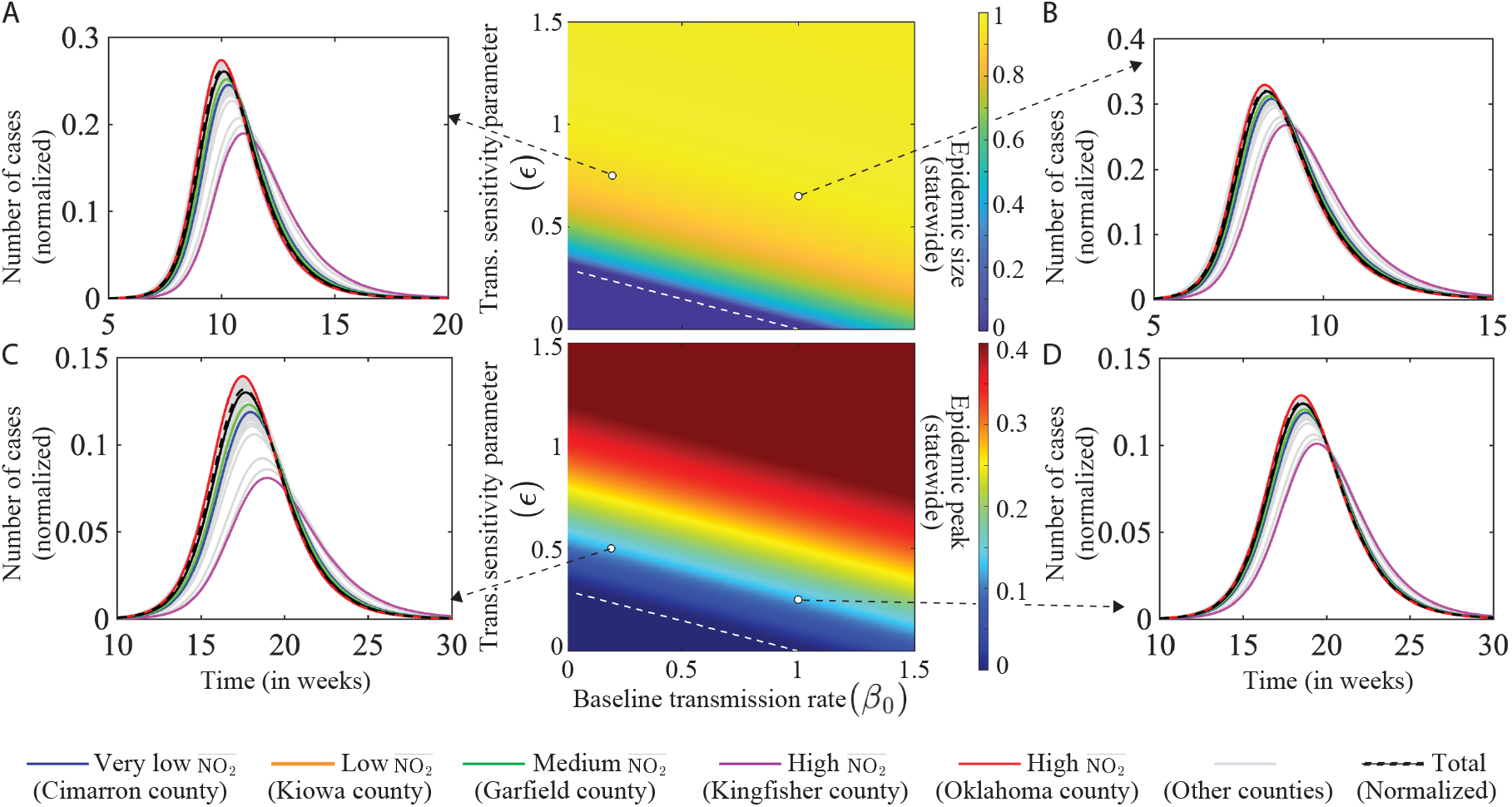
Sensitivity of epidemic outcomes to baseline transmission (*β*_0_) and NO_2_ sensitivity (*ϵ*). Epidemic outcomes vary with the baseline infection rate (*β*_0_, x-axis) and NO_2_-related transmission sensitivity (*ϵ*, y-axis). The central heatmaps illustrate the statewide epidemic size (top) and epidemic peak (bottom) as functions of the baseline infection rate (*β*_0_) and NO_2_ sensitivity (*ϵ*). White dashed lines represent the region where *R*_0_ = 1. Four representative parameter combinations are highlighted. **A, B**, High NO_2_ sensitivity leads to early, synchronized outbreaks across counties, largely independent of county-specific NO_2_ levels. **C, D**, Low NO_2_ sensitivity results in delayed and asynchronous county-level outbreaks, with very low NO_2_ counties (e.g., Cimarron) peaking substantially later than high NO_2_ counties (e.g., Oklahoma), highlighting the role of environmental sensitivity in shaping spatial heterogeneity in transmission. Higher baseline NO_2_ levels are associated with faster and more intense epidemic dynamics, whereas lower NO_2_ levels are associated with slower and delayed outbreaks. Parameters: **A** (*β*_0_, *ϵ*) = (0.2, 0.75), **B** (1, 0.65), **C** (0.2, 0.5), and **D** (1, 0.25). The recovery rate is fixed at *γ* = 1 (see Table 3 for other parameters).

### 4.3 Model fitting and performance comparison

We fitted our NO_2^-^_ and mobility-informed model to the first COVID-19 wave in Oklahoma (March 07, 2020 – July 03, 2021) and compared its county-level performance to a standard SIR model. Here, the NO_2^-^_ and mobility-informed model consistently achieved higher county-level *R*^2^ values (mean *R*^2^ = 0.663 vs. 0.556 for the SIR model) and lower nRMSE (mean = 0.126 vs. 0.146) and PAE (mean = 0.449 vs. 0.613) across all 77 counties, as shown in Fig. 3A. Supplementary Table S1 summarizes county-level *R*^2^, nRMSE, and PAE values computed across all 77 Oklahoma counties for both the SIR model and the NO_2^-^_ and mobility-informed model during the fitting of the first COVID-19 wave in Oklahoma. County-level win maps in Fig. 3B highlight the locations where the NO_2^-^_ and mobility-informed model outperformed the SIR model. Across the entire study period (March 07, 2020 – July 03, 2021), the NO_2^-^_ and mobility-informed model outperformed the SIR model in 76.62% of counties when evaluated using both nRMSE and *R*^2^. Additionally, it surpassed the SIR model in 85.71% of counties, as measured by the PAE, as shown in the Supplementary Table S2. The improved performance is illustrated in three selected time series for Creek, Rogers, and Cotton County (Fig. 3C), where the NO_2^-^_ and mobility-informed model more accurately tracks observed case counts, capturing peak timing and magnitude with narrower confidence intervals. This improvement is particularly striking in Cotton County, where the SIR model fails to explain any of the observed variance (*R*^2^ = −0.02 versus *R*^2^ = 0.79).

**Figure 3:**
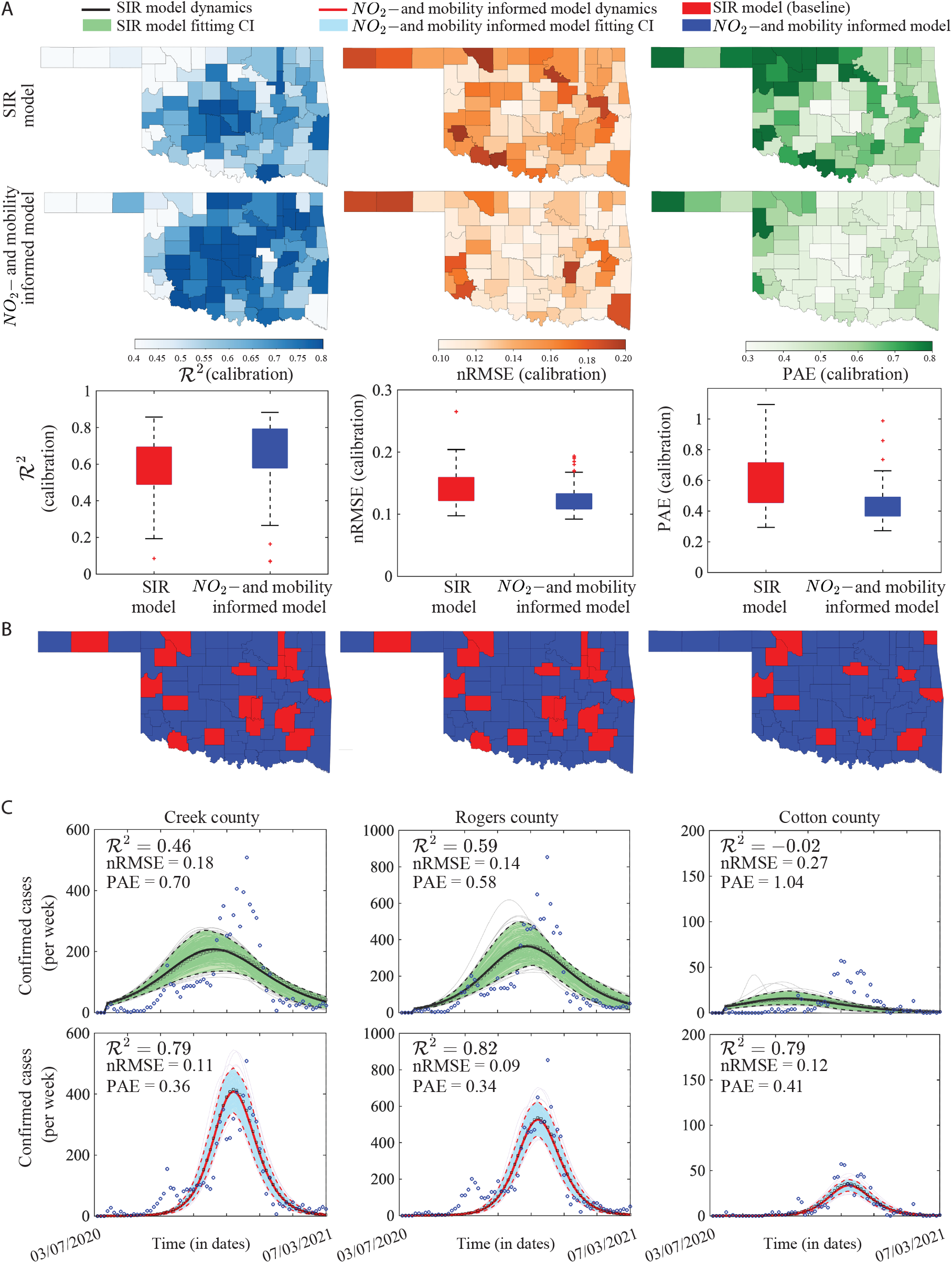
Model fits for the first COVID-19 wave in Oklahoma: NO_2^-^_ and mobility-informed model versus standard SIR model. **A**, County-level evaluation of model performance using three metrics: coefficient of determination (*R*^2^), normalized root mean square error (nRMSE), and percent absolute error (PAE) (definition provided in supplementary material B). Here, Oklahoma State’s county-level maps show spatial variation in model performance for the SIR model (top row) and the NO_2^-^_ and mobility-informed model (middle row). Across most counties, the informed model exhibits higher *R*^2^ values and lower nRMSE and PAE values. Box plots (bottom row) summarize metric distributions across all counties, showing higher median *R*^2^ and lower median nRMSE and PAE for the NO_2_ and mobility-informed model (red) compared with the SIR model (blue). **B**, County-level win maps showing counties where the NO_2^-^_ and mobility-informed model achieves better performance than the SIR model. **C**, Time-series fits for representative counties (Creek, Rogers, and Cotton) from March 07, 2020 to July 03, 2021. The NO_2_ and mobility-informed model (red line) more closely tracks observed confirmed cases (blue dots) than the SIR model (black line), yielding consistently higher *R*^2^ values. Shaded regions indicate model confidence intervals (green for SIR; light blue for the NO_2^-^_ and mobility-informed model).

### 4.4 Forecasting COVID-19 incidence across epidemic stages

We evaluated model forecasts across five different epidemic stages: Early, Surge, Peak, Post-Peak, and Decline (see Table 1 for specific dates). Fig. 4A depicts each stage in the time series of COVID-19 confirmed cases in Tulsa County, which was the first location in Oklahoma to report COVID cases. Here, we generated 1-, 2-, 3-, 4-, and 5-week-ahead predictions for each county across epidemic stages using both SIR and NO_2^-^_ and mobility-informed models, and reported the errors based on them. Forecasting accuracy was quantified using nRMSE (Fig. 4B-C) and PAE (Fig. 4D-E). Fig. 4B exhibits choropleth maps of Oklahoma with county-level performance in terms of nRMSE, along with boxplots, while win maps are depicted in Fig. 4C. Supplementary Tables S5 and S6 provide summary statistics of county-level nRMSE and PAE values, respectively, computed across all 77 Oklahoma counties for the SIR model and the NO_2^-^_ and mobility-informed model during different epidemic stages.

**Figure 4:**
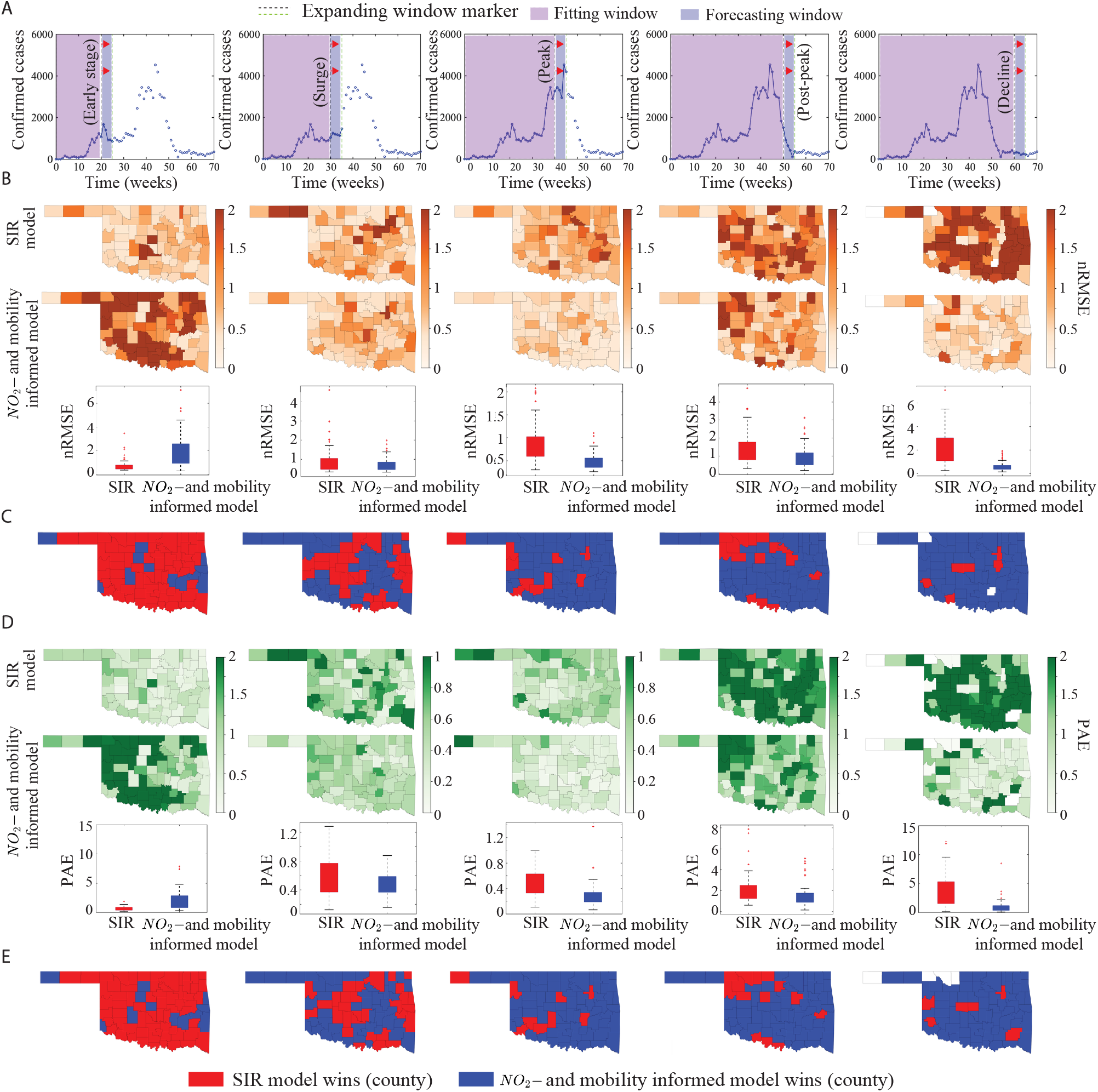
Forecasting performance of SIR and NO_2^-^_ and mobility-informed models across epidemic stages. **A**, Five-week-ahead predictions across epidemic stages (Early, Surge, Peak, Post-peak, Decline). **B**, County-level distributions of nRMSE shown as maps and box plots for each stage. Median nRMSE values for the NO_2^-^_ and mobility-informed model were lower than those of the SIR model during the surge (0.601 vs. 0.700), peak (0.437 vs. 0.793), post-peak (0.823 vs. 1.131), and decline (0.490 vs. 2.084) stages, indicating improved forecasting performance across most epidemic phases. (see details in Table S5 in Supplementary Material). **C**, County-level nRMSE win maps indicating which model achieves lower nRMSE. The NO_2^-^_ and mobility-informed model outperformed the SIR model in 59.74%, 87.01%, 80.52%, and 88.31% of counties during the surge, peak, post-peak, and decline stages, respectively, whereas the SIR model performed better during the early stage (84.42% of counties) (see details in Table S7 in Supplementary Material). **D**, Distributions of PAE across counties for each stage. Similarly, median PAE values for the NO_2^-^_ and mobility-informed model were lower than those of the SIR model during the surge (0.496 vs. 0.575), peak (0.254 vs. 0.508), post-peak (1.271 vs. 1.868), and decline (0.569 vs. 2.667) stages, indicating improved forecasting performance across most epidemic phases (see details in Table S6 in the Supplementary Material). **E**, County-level PAE win maps. Here, the NO_2^-^_ and mobility-informed model achieved lower PAE values in 61.04%, 88.31%, 81.82%, and 87.01% of counties during the surge, peak, post-peak, and decline stages, respectively, while the SIR model showed better performance during the early stage (85.71% of counties) (see details in Table S7 in the Supplementary Material). In panels C and E, red indicates counties where the SIR model performs better, blue indicates counties where the NO_2^-^_ and mobility-informed model performs better, and white indicates counties where neither model shows a clear advantage.

In the Early stage, the SIR model outperforms the NO_2^-^_ and mobility-informed model across most counties, with lower median nRMSE and a substantially tighter distribution. Two factors explain this pattern. First, the two models face a methodological asymmetry: the metapopulation model fits all 77 counties jointly starting from the first reported case in Tulsa on 7 March 2020, whereas the SIR model is fit independently in each county, starting from its own first nonzero reported case. For counties seeded later in the Early window, this yields a shorter time series well-described by the exponential growth phase of the standard SIR model, favoring the simpler fit. Second, case reporting in Oklahoma was substantially incomplete during the early months of the pandemic, particularly in rural counties with limited testing infrastructure. In this regime, the metapopulation model likely produces best-fit trajectories that predict case rises not reflected in the reported data, penalizing the model for capturing transmission that occurred but went undetected.

For the other epidemic stages, this pattern reversed decisively. The NO_2^-^_ and mobility-informed model outperformed the SIR model in 59.74% of counties during the Surge stage, and in 87.01%, 80.52%, and 88.31% during the Peak, Post-peak, and Decline stages, respectively. These results are consistent with those obtained using PAE (Fig. 4D-E and Supplementary Table S7). Beyond the increased win rate, the NO_2^-^_ and mobility-informed model’s error distributions tightened substantially across most of these stages (see Supplementary Tables S5 and S6 for standard deviation values). In contrast, the SIR model exhibited growing variability in later stages, with several counties showing nRMSE values that were two- to three-fold higher than the NO_2_ and mobility-informed model’s median. The advantage was most pronounced during the Peak and Post-peak stages, when spatial heterogeneity in transmission and mobility-driven coupling between counties were likely most relevant to capturing observed dynamics.

### 4.5 Model fits and forecasts across epidemic stages: time series from representative counties

We illustrate forecasting performance by presenting time series from five counties in distinct epidemic phases: Tulsa (Early), Oklahoma (Surge), Noble (Peak), Cherokee (Post-peak), and Latimer (Decline). These counties were selected as representative examples of distinct epidemic phases and spatial heterogeneity in outbreak dynamics across Oklahoma, capturing diverse temporal patterns in incidence rather than being chosen based on model performance metrics. Fig. 5A depicts training and forecasting windows for the early stages of the epidemic, along with a map of Oklahoma and selected counties. For the Early stage in Tulsa County (Fig. 5B), both models reasonably captured the uptrend and projected a rise in COVID-19 cases, while in reality, cases rose sharply but then declined until around week 30. Forecast values from the NO_2^-^_ and mobility-informed model were overall higher than those produced by the SIR model, resulting in decreased performance (see nRMSE values).

**Figure 5:**
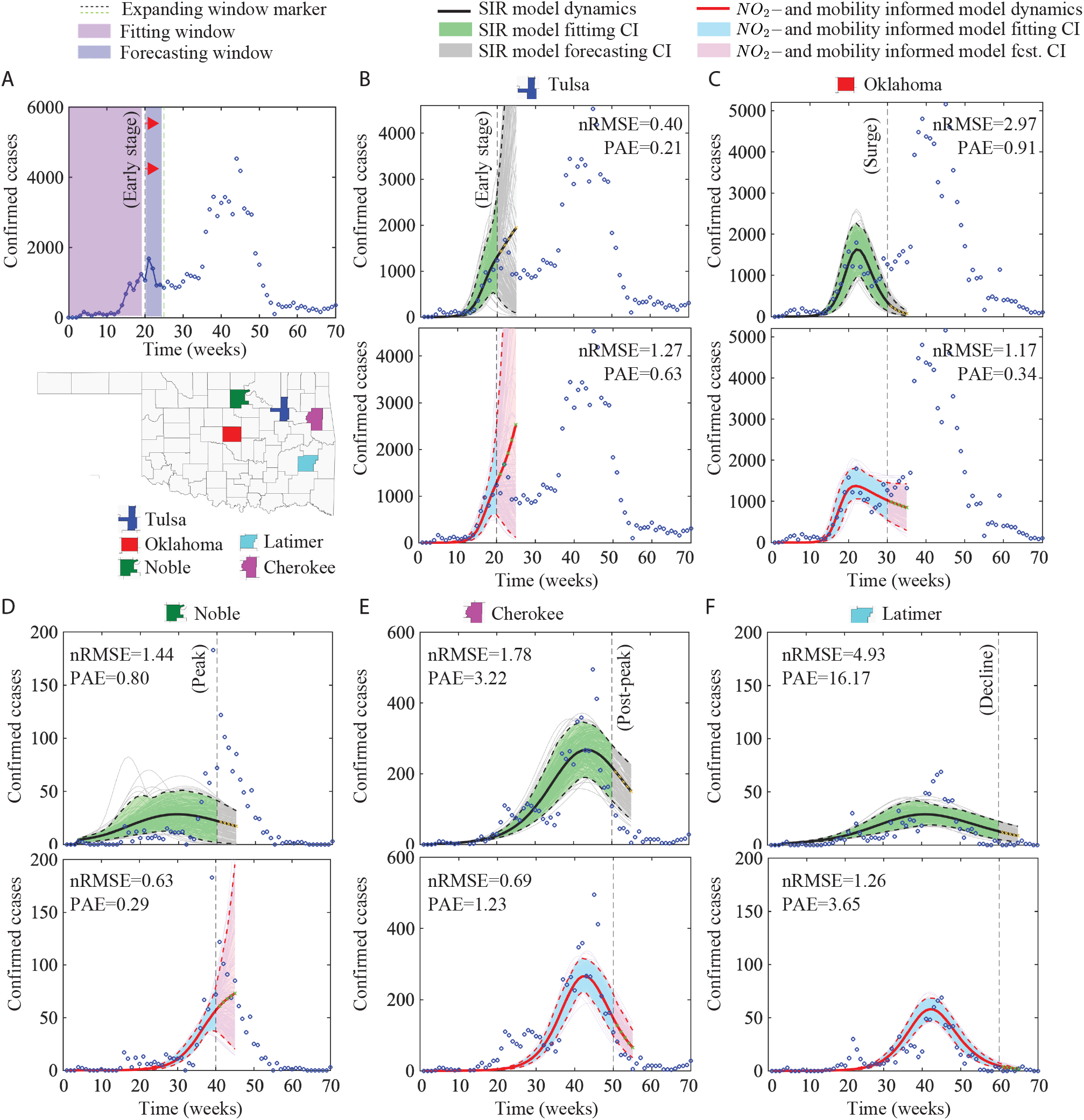
Representative county-level forecast across epidemic stages. **A**, Schematic of the Early stage window for model fitting (light purple shaded area) and five-week-ahead forecasting (light blue shaded area), alongside an Oklahoma county-level map and five representative locations: Tulsa, Oklahoma, Latimer, Noble, and Cherokee counties. **B–F**, County-specific forecasts for the five counties across different epidemic phases: Early (Tulsa, B), Surge (Oklahoma, C), Peak (Noble, D), Post-peak (Cherokee, E), and Decline (Latimer, F). Blue dots indicate observed confirmed cases. The SIR model (black solid line) with fitting confidence interval (light green) and forecasting confidence interval (light gray) fits the data well during the Early phase. In contrast, the NO_2^-^_ and mobility-informed model (red solid line) with fitting confidence interval (light blue) and forecasting confidence interval (light pink) provides more accurate fits and forecasts during later stages (Surge, Peak, Post-peak, and Decline).

At later stages, the scenario changed, and the time series demonstrate the impact of integrating NO_2_ and mobility data into a modeling pipeline. For the Surge stage in Oklahoma County, the SIR model captured a first surge but, as a consequence of the model structure, predicted a very low number of reported cases, while observed counts actually increased. In contrast, the NO_2^-^_ and mobility-informed model predicted a slight decrease in cases, also not capturing the increase in COVID-19 activity, but with predicted values much closer to the ground truth (Fig. 5C). The discrepancies between forecasts were even more dramatic at the Peak stage in Noble County (Fig. 5D), with the SIR model failing to fit the data and predicting a downtrend in cases instead of a peak. The NO_2^-^_ and mobility-informed model not only fit the 40-week training data but also predicted case counts reasonably well. For the Post-peak and Decline stages (Figs. 5E and F), model fits from the network model also improved in comparison to the SIR model, along with narrower forecasting confidence intervals and improved agreement with observed case counts. These representative examples reflect the county-level error metrics shown in Fig. 4, highlighting the improved predictive performance of the NO_2^-^_ and mobility-informed model in comparison to the SIR model across all epidemic stages except the Early stage.

### 4.6 Isolating the effect of NO_2_ on forecasting performance

To isolate the specific contribution of NO_2_-informed transmission heterogeneity from that of mobility-driven connectivity, we compared the NO_2^-^_ and mobility-informed model against a reduced model that retains the same mobility structure but excludes NO_2_-dependent transmission heterogeneity (i.e., we set *ϵ* = 0). The NO_2^-^_ and mobility-informed model is fitted under the constraint *ϵ* ≥ 0.01, reflecting the hypothesis that NO_2_ exposure increases the per-contact probability of transmission. Fig. 6 shows county-level differences in forecasting error (ΔnRMSE and ΔPAE) between the two models across the five epidemic stages. Additionally, supplementary Tables S8, S9 and S10 report the corresponding county-level predictive and win percentages. Consistent with the SIR comparison (Fig. 4), the NO_2^-^_ and mobility-informed model underperforms the mobility-informed model during the Early stage (14.29% nRMSE, 15.58% PAE win rate) but wins in the clear majority of counties during Surge (94.81% nRMSE, 90.91% PAE) and Post-peak (88.31% nRMSE, 85.71% PAE), and in a smaller majority during Peak (62.34% nRMSE, 61.04% PAE). Notably, this advantage does not persist into the Decline stage, where the mobility-informed model again wins the majority of counties (63.63% nRMSE, 66.23% PAE), mirroring the reduced relative performance observed in the Early stage. The results indicate that the improvements over the SIR model reported above are not attributable to mobility alone: NO_2_-dependent transmission heterogeneity provides an additional, spatially consistent forecasting benefit specifically during the Surge, Peak, and Post-peak stages, when the epidemic is accelerating or turning over, but offers no added benefit once cases are already receding statewide. We also compared the mobility-informed model and the NO_2^-^_ and mobility-informed model over the full study period, as done in Section 4.3 for the SIR comparison. Both models achieve nearly identical in-sample error across all 77 counties, with matching mean and median values of *R*^2^, nRMSE, and PAE (Supplementary Table S3); the mobility-informed model wins a modest majority of counties (64% by nRMSE and *R*^2^, 55% by PAE, Supplementary Table S4).

**Figure 6:**
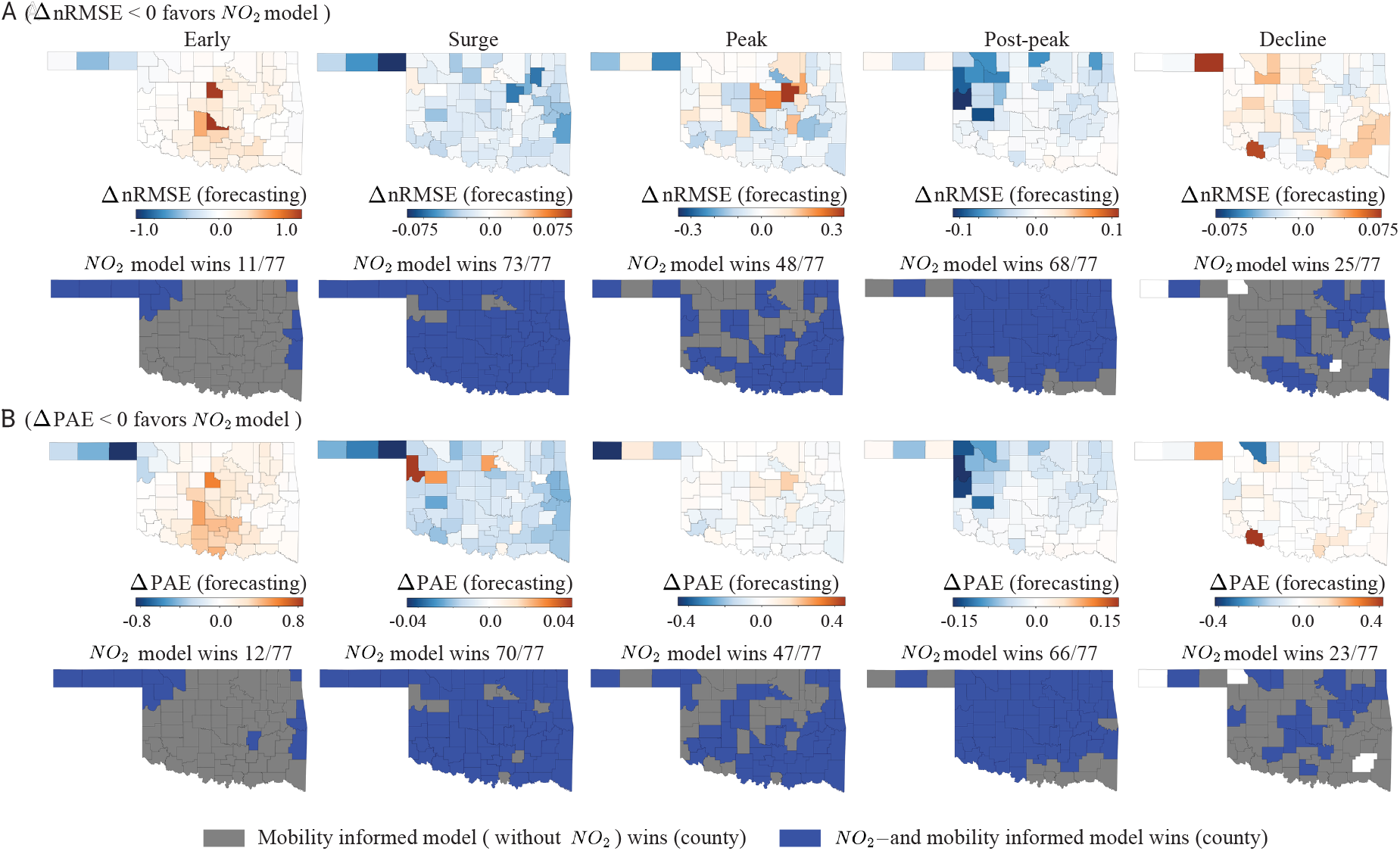
County-level spatial heterogeneity in forecasting performance between the NO_2^-^_ and mobility-informed model and the mobility-informed model (without NO_2_) across epidemic stages. **A**, County-level maps of ΔnRMSE (forecasting), calculated as the difference in nRMSE between the two models, for the Early, Surge, Peak, Post-peak, and Decline stages (ΔnRMSE *<* 0 favours the NO_2_-informed model (with NO_2_), blue shading). Below each map, the corresponding win/loss map classifies each of the 77 Oklahoma counties by which model achieves lower forecasting error: the NO_2^-^_ and mobility-informed model (blue), the mobility-informed model without NO_2_ (grey), or white indicates counties (total 3) where neither model shows a clear advantage. Consistent with the representative time series in Fig. 5, the NO_2^-^_ and mobility-informed model underperforms during the Early and Decline stages, winning in 11/77 and 25/77 counties, respectively, but shows a marked improvement, winning in 73/77 (Surge), 48/77 (Peak), and 68/77 (Post-peak). **B**, Corresponding county-level maps of ΔPAE (forecasting) and associated win/loss classifications (ΔPAE *<* 0 favours the NO_2_-informed model), showing a similar pattern: the NO_2^-^_ and mobility-informed model wins in 12/77 (Early), 70/77 (Surge), 47/77 (Peak), 66/77 (Post-peak), and 23/77 (Decline) counties.

## 5. Discussion

The COVID-19 pandemic revealed the need for epidemic models that capture the heterogeneity shaping early outbreak dynamics. Among drivers of such heterogeneity, the effects of chemical exposures, and in particular air pollution, have remained largely understudied in mechanistic transmission modeling. Here we developed a NO_2^-^_ and mobility-informed metapopulation model that incorporates county-level nitrogen dioxide concentrations directly into county-specific infection rates across the 77 counties of Oklahoma. When applied to the first COVID-19 wave in the state, the model improved the characterization of county-level dynamics relative to the SIR model across the fitting period and most epidemic stages.

Our model links NO_2_ to infection rates through a first-order (linear) term. Using numerical simulations, we showed that when such rates are highly sensitive to NO_2_ (i.e., higher *ϵ*), early/synchronized outbreaks are observed, whereas lower sensitivity generated delayed and asynchronous trajectories across counties (Fig. 2).

When fitting the model to the first COVID-19 wave in Oklahoma (Fig. 3), the NO_2^-^_ and mobility-informed model outperformed the SIR model in 76.62% of counties based on both nRMSE and *R*^2^, and in 85.71% of counties based on PAE (Supplementary Table S2). The improvement was particularly striking in counties such as Cotton, where the SIR model failed to explain any of the observed variance (*R*^2^ = −0.02) while the NO_2^-^_ and mobility-informed model achieved *R*^2^ = 0.79. The model also exhibited improved forecasting performance relative to the SIR model across most epidemic stages (Fig. 4). While the standard SIR model performed reasonably well during the Early phase, its performance deteriorated during the Surge, Peak, Post-peak, and Decline phases. This decline in performance is expected: as the epidemic matures, the classic SIR model – which ignores spatial structure and treats each county as an isolated node – tends to misrepresent local dynamics and progressively lose predictive fidelity (*61*). In contrast, the NO_2^-^_ and mobility-informed model maintained substantially lower forecasting error across later epidemic stages, achieving county-level win rates of 59.74% (Surge), 87.01% (Peak), 80.52% (Post-peak), and 88.31% (Decline) in terms of nRMSE (Supplementary Table S7). Representative county-level forecasts (Fig. 5) further confirmed that the model more accurately captured both the timing and magnitude of epidemic peaks during these later phases.

The NO_2^-^_ and mobility-informed model differs from the SIR model in both its network structure and its environmental transmission term; hence, an important question is how much of the reported improvement is attributable specifically to NO_2_ rather than to mobility-driven connectivity alone. Comparing our full model against a reduced model that retains the same mobility structure but excludes NO_2_-dependent transmission heterogeneity showed that the NO_2_ term itself contributes meaningfully to forecasting accuracy: the NO_2^-^_ and mobility-informed model outperformed the mobility-only model in the majority of counties during the Surge, Peak, and Post-peak stages (Fig. 6). This advantage did not extend to the Early and Decline stages, where the mobility-only model instead won the majority of counties, mirroring the reduced relative performance of the NO_2^-^_ and mobility-informed model against the SIR model during the Early stage. This indicates that the forecasting improvements reported above are not simply a byproduct of incorporating spatial connectivity, and that environmental exposure carries forecasting-relevant information beyond what mobility patterns alone provide.

Some limitations of the current study warrant acknowledgment. The relationship between NO_2_ exposure and transmission was modeled as a linear function of county-level concentrations. The underlying biological mechanisms are likely more complex, and nonlinear formulations capturing threshold or saturation effects could also be adopted. The SIR structure adopted here does not account for loss of immunity and vaccination, factors that became increasingly relevant beyond the first COVID-19 wave. Another limitation concerns the interpretation of the NO_2_ term in Eq. 3. Our choice to parameterize infection rates by county-level NO_2_ concentrations rests on biological mechanisms in which chronic NO_2_ exposure impairs respiratory defenses and alters immune function, plausibly raising the probability that a contact results in infection. Since *β*_*j*_ depends on ⟨NO_2_⟩_*j*_ through a monotone transformation, any county-level covariate strongly correlated with NO_2_ (urbanicity indices or population density, for instance) could induce a similar ordering of county-specific infection rates and could yield comparable fitting and forecasting performance. Distinguishing NO_2_ from such correlates would require comparing models parameterized by alternative covariates.

This study demonstrated that air pollution data can be incorporated directly into the transmission structure of a mechanistic epidemic model. When applied to the first COVID-19 wave in Oklahoma, the NO_2^-^_ and mobility-informed model improved the characterization of county-level dynamics relative to the SIR model, and the NO_2_ term itself carried forecasting-relevant information beyond that provided by mobility during the stages of high incidence. The formulation is not specific to NO_2_ or to SARS-CoV-2: any exposure that plausibly modulates the probability of transmission per contact and varies across the units of a metapopulation can enter an infection rate in the same way, making the approach extensible to other pollutants, such as particulate matter, and to other respiratory pathogens. As satellite-derived environmental data are spatially resolved and routinely updated, air pollutants offer an accessible source of structured heterogeneity for spatially explicit forecasting. In this sense, our findings demonstrate that mechanistic models of emerging pathogens may benefit from representing the environmental conditions in which transmission occurs.

## Data Availability

All data produced in the present study are available upon reasonable request to the authors.

## Acknowledgments

This research was supported by the National Science Foundation under grant NSF DMS 2327844 (IHBEM: Using socioeconomic, behavioral, and environmental data to understand disease dynamics — exploring COVID-19 outcomes in Oklahoma).

## Supplementary Material

This Appendix provides additional technical details on data sources, model calibration procedures, uncertainty quantification, and evaluation metrics used in the main text.

### A Additional Details on NO_2_ Data and Model Calibration

This section provides additional details on the NO_2_ datasets and parameter-estimation procedures used for model calibration. We first describe the environmental NO_2_ dataset used to construct spatially varying infection rates.

#### A.1 Nitrogen dioxide (NO_2_) data: technical details

Following (*48*), county-level NO_2_ data were obtained from the Sentinel-5P TROPOMI instrument, spanning January 1, 2020, through December 31, 2023, and covering all U.S. counties identified by state and county FIPS codes. Daily measurements were aggregated at the county level and expressed in mg/m^3^. Observations were retained only if the quality assurance (QA) value exceeded standard thresholds. For each county *j*, the mean NO_2_ concentration ⟨NO_2_⟩_*j*_ was computed over the pre-pandemic period (January 1 – March 1, 2020), ranging from 2.2 to 3.6 mg/m^3^ across the 77 Oklahoma counties (Fig. 1B). Pre-pandemic concentrations (January 1–March 1, 2020) served as a baseline, while pandemic averages (January–December 2020) ranged from 2.835 to 3.62 mg/m^3^ across the 77 Oklahoma counties. These NO_2_ data were subsequently combined with mobility information to simulate environmentally modulated transmission dynamics in our epidemiological model. We next detail the optimization procedures used to estimate model parameters for both the SIR and network models.

#### A.2 Optimization procedure for SIR model fitting

The SIR model includes three unknown parameters: the infection rate *β*, the recovery rate *γ*, and the reporting rate *ρ*. To reduce sensitivity to initial guesses and avoid convergence to local minima, we employed a multistart randomized optimization strategy. For each optimization run, an initial parameter vector was generated randomly within predefined bounds: 0.1 ≤ *β* ≤ 3, 0.001 ≤ *ρ* ≤ 0.2, 0.5 ≤ *γ* ≤ 1.

Specifically, the initial parameter vector was sampled as

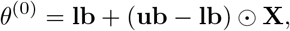

where *θ*^(0)^ = (*β*^(0)^, *ρ*^(0)^, *γ*^(0)^) denotes the initial parameter vector, **lb** and **ub** represent the lower and upper parameter bounds, respectively, **X** ∼ *U* (0, 1)^3^ is a vector of independent uniform random variables, and ⊙ denotes element-wise multiplication.

Then, the sampled initial values were used as starting points for the constrained nonlinear optimization routine (fmincon) to minimize the sum of squared residuals (SSR) between observed and model-predicted cases (Eq. 6). For each county, the optimization was repeated across 200 random initializations (200 iterations) to improve robustness and reduce the risk of local minima. This procedure was repeated independently for each county and for all expanding training windows described in Table 1. This approach produced robust county-specific estimates of *β, γ*, and *ρ*, capturing spatial heterogeneity in epidemic dynamics.

#### A.3 Optimization procedure for NO_2^-^_ and mobility-informed model fitting

The network model includes four unknown parameters, *θ* = (*β*_0_, *ϵ, γ, ρ*). To reduce sensitivity to initial guesses and avoid convergence to local minima, we employed a multi-start randomized optimization strategy. For each optimization run, an initial parameter vector was generated randomly within predefined bounds: 0.01 ≤ *β*_0_ ≤ 0.25 + max(NO2_stats_table.Mean_NO2), 0.01 ≤ *ϵ* ≤ 1, 0.0001 ≤ *γ* ≤ 2, 0.0001 ≤ *ρ* ≤ 0.4.

Here, the infection rate *β*_0_ was bounded between 0.01 and 0.25+max(NO_2__stats_table.Mean_NO2), where the upper bound accounts for the maximum observed NO_2_ concentration. The NO_2_ scaling parameter *ϵ*, recovery rate *γ*, and reporting rate *ρ* were bounded as indicated above.

Specifically, the initial parameter vector was sampled as

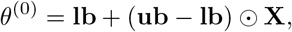

where *θ*^(0)^ = (*β*^(0)^, *ϵ*^(0)^, *γ*^(0)^, *ρ*^(0)^) denotes the initial parameter vector, **lb** and **ub** represent the lower and upper parameter bounds, respectively, **X** ∼ *U* (0, 1)^4^ is a vector of independent uniform random variables, and ⊙ denotes element-wise multiplication.

Then, the sampled initial values were used as starting points for the constrained nonlinear optimization routine (fmincon) to minimize the sum of squared residuals (SSR) between observed and model-predicted cases (Eq. 6). The optimization was repeated 200 times with random initializations to improve robustness and reduce the risk of convergence to local minima. Unlike the SIR model, which is fitted independently in each county, the NO_2^-^_ and mobility-informed model was fitted jointly across all 77 counties, minimizing the total sum of squared residuals over the network. We repeated this procedure for all expanding training windows described in Table 1, producing a single set of global estimates of *β*_0_, *ϵ, γ*, and *ρ* per window, with spatial heterogeneity in infection rates arising from ⟨NO_2_⟩_*j*_ through Eq. 3.

We now describe the parametric bootstrap procedures used to quantify uncertainty in model estimates and forecasts.

##### Algorithm S1

Parametric Bootstrap for SIR Model Uncertainty

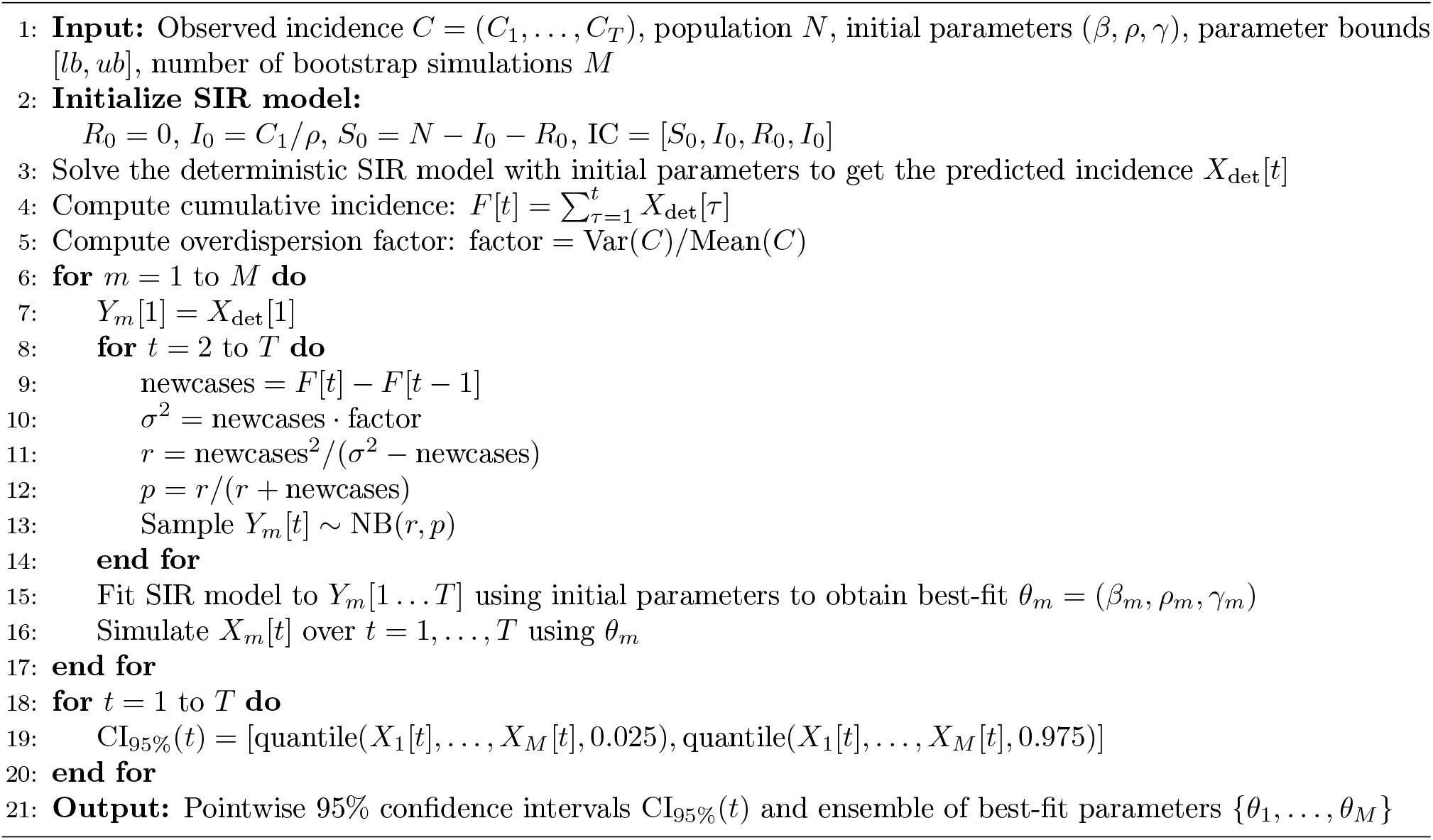

##### Algorithm S2

Parametric Bootstrap for NO_2^-^_ and mobility-informed model Uncertainty and Forecasting

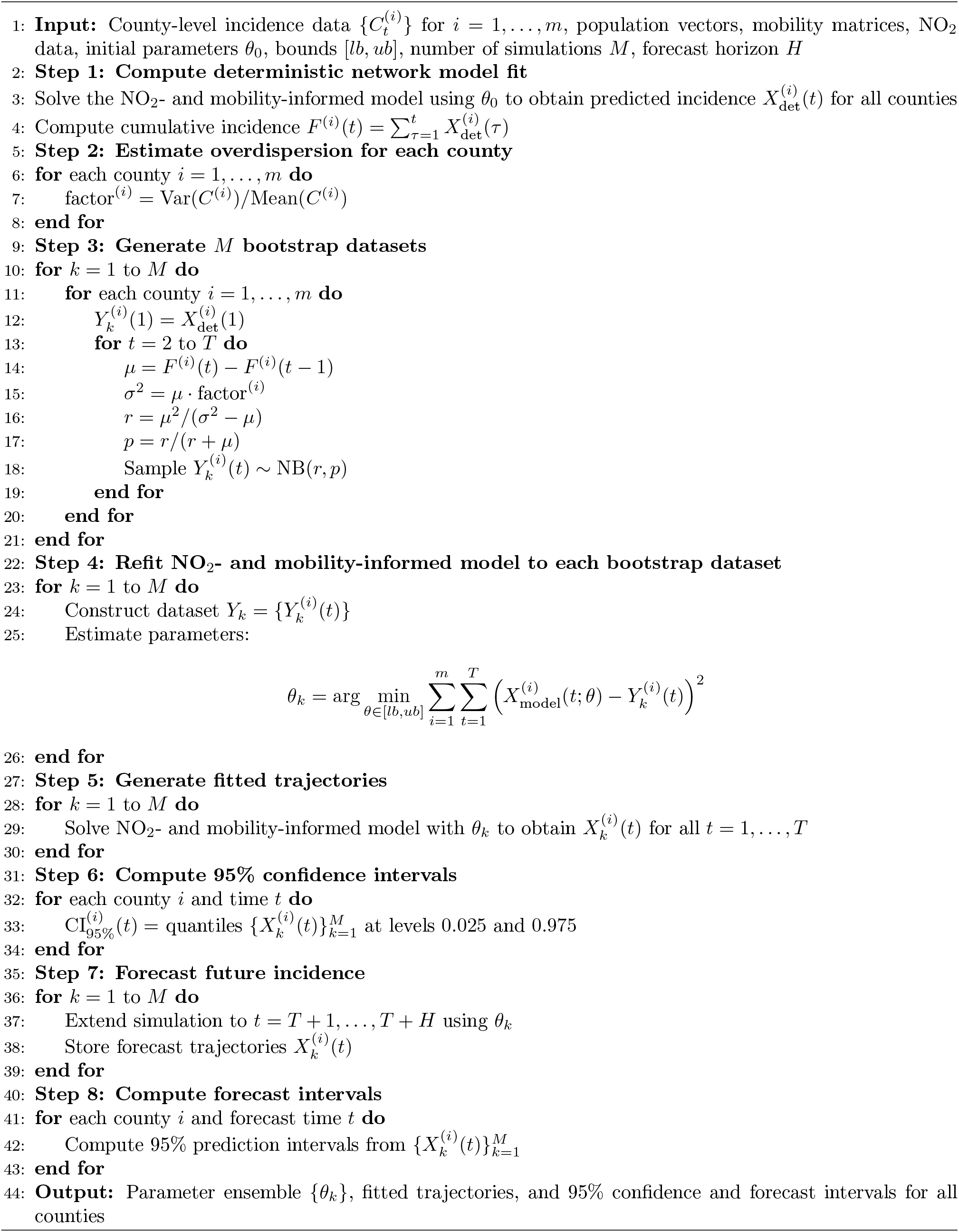

### B. Fitting and forecasting evaluation metrics

To evaluate and compare the fitting and forecasting performance of the NO_2^-^_ and mobility-informed model with the SIR model, we employed several standard goodness-of-fit and forecasting accuracy metrics, including the *R*^2^, nRMSE, and PAE. These metrics quantify complementary aspects of model performance and forecasting accuracy across counties and epidemic stages.

#### Coefficient of Determination (*R*^2^)

The coefficient of determination *R*^2^ measures the proportion of variance in the observed data that is explained by the model. Let *y*_1_, …, *y*_*T*_ be the observed values and *ŷ*_1_, …, *ŷ*_*T*_ the corresponding model predictions. Then, *R*^2^ is given by:

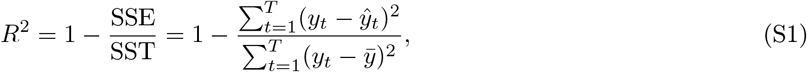

where 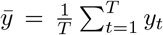 is the mean of the observed values. An *R*^2^ value close to 1 indicates that the model explains most of the variance in the observed data, whereas values near 0 indicate poor explanatory power. Negative values may occur when the model performs worse than simply predicting the mean.

#### Normalized root mean squared error

The root mean squared error (RMSE) is defined as the square root of the average squared differences between the predicted and actual values. Additionally, the normalized root mean square error (nRMSE) measures the magnitude of prediction errors in relation to the range of the actual data.

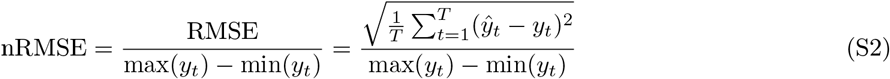

The RMSE metric emphasizes larger errors more strongly than MAE and provides insight into the typical magnitude of prediction errors. For example, when the nRMSE values range from 0 to 1, a value near 0 indicates that the predictions closely match the actual data. As the value increases toward 1, the model becomes less accurate. Additionally, values approaching 1 represent significant prediction errors and indicate that the model is performing poorly.

#### Percent Absolute Error

We define percent absolute error (PAE) as the ratio of the mean absolute error to the mean of the observed monthly case counts over the evaluation period. Let *y*_1_, …, *y*_*T*_ be the ground-truth reported case counts, and *ŷ*_1_, …, *ŷ*_*T*_ be the corresponding predicted case counts. Then, the PAE is given by:

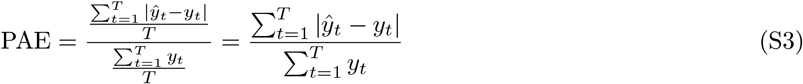

Using these evaluation metrics, we quantified and compared the county-level fitting and forecasting performance of the SIR model and the NO_2^-^_ and mobility-informed model across all 77 Oklahoma counties during the first COVID-19 wave.

#### B.1 Model performance: full fitting window

This subsection reports overall model fitting performance across the full study period (March 07, 2020 – July 03, 2021).

##### B.1.1 SIR model versus NO_2^-^_ and mobility-informed model

Table S1 summarizes county-level *R*^2^, nRMSE, and PAE statistics, and Table S2 reports the corresponding county-level win percentages for the SIR model and the NO_2^-^_ and mobility-informed model.

**Table S1:** Summary statistics for county-level *R*^2^, nRMSE, and PAE values were calculated across all 77 counties in Oklahoma for the SIR model, as well as for the NO_2^-^_ and mobility-informed model, during the model fitting of the first COVID-19 wave in Oklahoma.

| Metric | SIR Model |  |  | NO<sub>2</sub>- and Mobility-Informed Model |  |  |
| --- | --- | --- | --- | --- | --- | --- |
|  | Mean | Median | SD | Mean | Median | SD |
| $R^2$ | 0.556 | 0.587 | 0.198 | 0.663 | 0.714 | 0.178 |
| nRMSE | 0.146 | 0.144 | 0.031 | 0.126 | 0.119 | 0.025 |
| PAE | 0.613 | 0.596 | 0.197 | 0.449 | 0.421 | 0.121 |

**Table S2:** Full study period fitting performance (March 07, 2020 to July 03, 2021) for SIR model win (%) versus NO_2^-^_ and mobility-informed model win (%).

| Metric | SIR model win (%) | NO<sub>2</sub>- and mobility-informed model win (%) | Neither model win (%) |
| --- | --- | --- | --- |
| nRMSE | 23.38 | 76.62 | 0.00 |
| $R^2$ | 23.38 | 76.62 | 0.00 |
| PAE | 14.29 | 85.71 | 0.00 |

##### B.1.2 Mobility-informed model versus NO_2^-^_ and mobility-informed model

Table S3 and Table S4 report the corresponding full-window fitting comparison between the mobility-informed model (without NO_2_) and the NO_2^-^_ and mobility-informed model. Unlike the SIR comparison above (Tables S1 S2), the two models show nearly identical aggregate performance across all 77 counties, with essentially matching mean, median, and SD values for *R*^2^, nRMSE, and PAE, and the mobility-informed model winning a modest majority of counties (64% by nRMSE and *R*^2^, 55% by PAE).

**Table S3:** Summary statistics for county-level *R*^2^, nRMSE, and PAE values were calculated across all 77 counties in Oklahoma for the mobility-informed model, as well as for the NO_2^-^_ and mobility-informed model, during the model fitting of the first COVID-19 wave in Oklahoma.

| Metric | Mobility-informed model |  |  | NO<sub>2</sub>- and Mobility-Informed Model |  |  |
| --- | --- | --- | --- | --- | --- | --- |
|  | Mean | Median | SD | Mean | Median | SD |
| $R^2$ | 0.663 | 0.716 | 0.178 | 0.663 | 0.714 | 0.178 |
| nRMSE | 0.126 | 0.119 | 0.025 | 0.126 | 0.119 | 0.025 |
| PAE | 0.449 | 0.421 | 0.122 | 0.449 | 0.421 | 0.121 |

**Table S4:** Full study period fitting performance (March 07, 2020 to July 03, 2021) for mobility-informed model win (%) versus NO_2^-^_ and mobility-informed model win (%).

| Metric | Mobility-informed<br>model win (%) | NO<sub>2</sub>- and mobility-informed<br>model win (%) | Neither<br>model win (%) |
| --- | --- | --- | --- |
| nRMSE | 63.64 | 36.36 | 0.00 |
| $R^2$ | 63.64 | 36.36 | 0.00 |
| PAE | 54.55 | 45.45 | 0.00 |

#### B.2 Model performance at different epidemic stages

This subsection reports forecasting performance across the five epidemic stages (Early, Surge, Peak, Post-peak, Decline; see Table 1 for exact date ranges).

##### B.2.1 SIR model versus NO_2^-^_ and mobility-informed model

To evaluate the performance of the SIR model and the NO_2^-^_ and mobility-informed model across different epidemic stages in all 77 Oklahoma counties, we present a summary of the county-level nRMSE and PAE statistics in Tables S5 and S6, respectively. Additionally, Table S7 displays the county-level win percentages for each model, metric, and stage across all 77 counties.

**Table S5:** Summary statistics of county-level *nRMSE values* computed across all 77 Oklahoma counties for the SIR model and the NO_2^-^_ and mobility-informed model across epidemic stages. Here, the lower values indicate better predictive performance. Note that non-finite values such as NaN and Inf were ignored to calculate these results.

| Window | SIR Model |  |  | NO<sub>2</sub>- and Mobility-Informed Model |  |  |
| --- | --- | --- | --- | --- | --- | --- |
|  | Mean | Median | SD | Mean | Median | SD |
| Early | 0.702 | 0.586 | 0.478 | 1.959 | 1.572 | 1.392 |
| Surge | 0.890 | 0.700 | 0.665 | 0.720 | 0.601 | 0.342 |
| Peak | 0.875 | 0.793 | 0.408 | 0.483 | 0.437 | 0.177 |
| Post-peak | 1.412 | 1.131 | 0.942 | 1.004 | 0.823 | 0.680 |
| Decline | 2.218 | 2.084 | 1.434 | 0.580 | 0.490 | 0.349 |

**Table S6:** Summary statistics of county-level *PAE values* computed across all 77 Oklahoma counties for the SIR model and the NO_2^-^_ and mobility-informed model across epidemic stages. Here, the lower values indicate improved predictive performance. Note that non-finite values such as NaN and Inf were ignored to calculate these results.

| Window | SIR Model |  |  | NO<sub>2</sub>- and Mobility-Informed Model |  |  |
| --- | --- | --- | --- | --- | --- | --- |
|  | Mean | Median | SD | Mean | Median | SD |
| Early | 0.649 | 0.620 | 0.318 | 2.076 | 1.507 | 1.619 |
| Surge | 0.588 | 0.575 | 0.280 | 0.478 | 0.496 | 0.155 |
| Peak | 0.499 | 0.508 | 0.196 | 0.288 | 0.254 | 0.184 |
| Post-peak | 2.105 | 1.868 | 1.319 | 1.523 | 1.271 | 1.057 |
| Decline | 3.478 | 2.667 | 6.417 | 0.618 | 0.569 | 2.122 |

**Table S7:** County-level winning percentages comparing the SIR model and the NO_2^-^_ and mobility-informed model across epidemic stages and forecasting metrics. Percentages represent the proportion of total Oklahoma counties (77) for which each model achieved lower forecasting error or better predictive performance. “Neither” indicates counties where neither model demonstrated a clear performance advantage.

| Epidemic Stage | Metric | SIR model win (%) | NO<sub>2</sub>- and mobility-informed model win (%) | Neither model win (%) |
| --- | --- | --- | --- | --- |
| Early | nRMSE | 84.42 | 15.58 | 0.00 |
|  | PAE | 85.71 | 14.29 | 0.00 |
| Surge | nRMSE | 40.26 | 59.74 | 0.00 |
|  | PAE | 38.96 | 61.04 | 0.00 |
| Peak | nRMSE | 12.99 | 87.01 | 0.00 |
|  | PAE | 11.69 | 88.31 | 0.00 |
| Post-peak | nRMSE | 19.48 | 80.52 | 0.00 |
|  | PAE | 18.18 | 81.82 | 0.00 |
| Decline | nRMSE | 7.79 | 88.31 | 3.90 |
|  | PAE | 7.79 | 87.01 | 5.19 |

##### B.2.2 Mobility-informed model versus NO_2^-^_ and mobility-informed model

The following table reports the corresponding win percentages for the comparison isolating the contribution of NO_2_: the mobility-informed model without NO_2_ (*ϵ* = 0) versus the full NO_2^-^_ and mobility-informed model (see Section 4.6 and Fig. 6). To evaluate the performance of the mobility-informed model and the NO_2^-^_ and mobility-informed model across different epidemic stages in all 77 Oklahoma counties, we present a summary of the county-level nRMSE and PAE statistics in Tables S8 and S9, respectively. Additionally, Table S10 displays the county-level win percentages for each model, metric, and stage across all 77 counties.

**Table S8:** Summary statistics of county-level *nRMSE values* computed across all 77 Oklahoma counties for the mobility-informed model and the NO_2^-^_ and mobility-informed model across epidemic stages. Here, the lower values indicate better predictive performance. Note that non-finite values such as NaN and Inf were ignored to calculate these results.

| Window | Mobility-informed model |  |  | NO<sub>2</sub>- and Mobility-Informed Model |  |  |
| --- | --- | --- | --- | --- | --- | --- |
|  | Mean | Median | SD | Mean | Median | SD |
| Early | 1.811 | 1.423 | 1.251 | 1.959 | 1.572 | 1.392 |
| Surge | 0.741 | 0.611 | 0.351 | 0.720 | 0.601 | 0.342 |
| Peak | 0.493 | 0.430 | 0.156 | 0.483 | 0.437 | 0.177 |
| Post-peak | 1.023 | 0.844 | 0.700 | 1.004 | 0.823 | 0.680 |
| Decline | 0.571 | 0.497 | 0.336 | 0.580 | 0.490 | 0.349 |

**Table S9:**
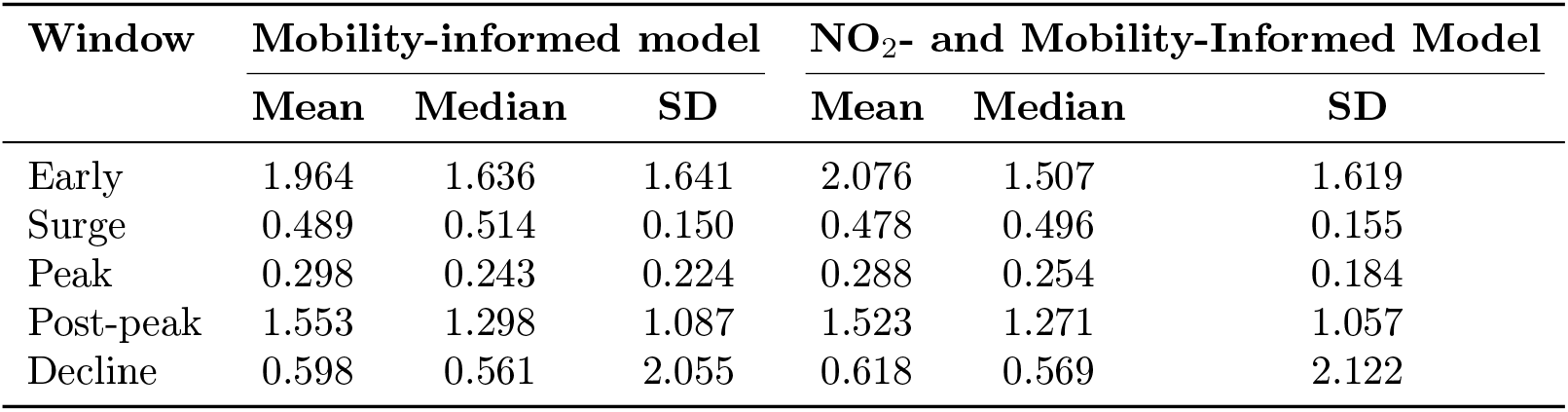
Summary statistics of county-level *PAE values* computed across all 77 Oklahoma counties for the mobility-informed model and the NO_2^-^_ and mobility-informed model across epidemic stages. Here, the lower values indicate improved predictive performance. Note that non-finite values such as NaN and Inf were ignored to calculate these results.

| Window | Mobility-informed model |  |  | NO<sub>2</sub>- and Mobility-Informed Model |  |  |
| --- | --- | --- | --- | --- | --- | --- |
|  | Mean | Median | SD | Mean | Median | SD |
| Early | 1.964 | 1.636 | 1.641 | 2.076 | 1.507 | 1.619 |
| Surge | 0.489 | 0.514 | 0.150 | 0.478 | 0.496 | 0.155 |
| Peak | 0.298 | 0.243 | 0.224 | 0.288 | 0.254 | 0.184 |
| Post-peak | 1.553 | 1.298 | 1.087 | 1.523 | 1.271 | 1.057 |
| Decline | 0.598 | 0.561 | 2.055 | 0.618 | 0.569 | 2.122 |

**Table S10:** County-level winning percentages comparing the mobility-informed model and the NO_2^-^_ and mobility-informed model across epidemic stages and forecasting metrics. Percentages represent the proportion of total Oklahoma counties (77) for which each model achieved lower forecasting error or better predictive performance. “Neither” indicates counties where neither model demonstrated a clear performance advantage.

| Epidemic Stage | Metric | Mobility-informed model win (%) | NO<sub>2</sub>- and mobility-informed model win (%) | Neither model win (%) |
| --- | --- | --- | --- | --- |
| Early | nRMSE | 85.71 | 14.29 | 0.00 |
|  | PAE | 84.42 | 15.58 | 0.00 |
| Surge | nRMSE | 5.19 | 94.81 | 0.00 |
|  | PAE | 9.09 | 90.91 | 0.00 |
| Peak | nRMSE | 37.66 | 62.34 | 0.00 |
|  | PAE | 38.96 | 61.04 | 0.00 |
| Post-peak | nRMSE | 11.69 | 88.31 | 0.00 |
|  | PAE | 14.29 | 85.71 | 0.00 |
| Decline | nRMSE | 63.63 | 32.47 | 3.90 |
|  | PAE | 66.23 | 29.87 | 3.90 |

## Notes

### Competing Interest Statement

The authors have declared no competing interest.

